# Comprehensive phenotyping of 3q29 deletion syndrome: recommendations for clinical care

**DOI:** 10.1101/2020.07.25.20162149

**Authors:** Rossana Sanchez Russo, Michael J. Gambello, Melissa M. Murphy, Katrina Aberizk, Emily Black, T. Lindsey Burrell, Grace Carlock, Joseph F. Cubells, Michael T. Epstein, Roberto Espana, Katrina Goines, Ryan Guest, Cheryl Klaiman, Sookyong Koh, Elizabeth Leslie, Longchuan Li, Derek Novacek, Celine A. Saulnier, Esra Sefik, Sarah Schultz, Elaine Walker, Stormi White, The Emory 3q29 Project, Jennifer Gladys Mulle

## Abstract

**Purpose:** To understand the consequences of the 3q29 deletion on medical, neurodevelopmental, psychiatric, and neurological sequalae by systematic evaluation of affected individuals. To develop evidence-based recommendations using these data for effective clinical care.

**Methods:** 32 Individuals with the 3q29 deletion were evaluated using a defined phenotyping protocol and standardized data collection instruments.

**Results:** Medical manifestations were varied and reported across nearly every bodily system, with congenital heart defects (25%) the most severe and heterogeneous gastrointestinal symptoms (81%) the most common. Physical exam revealed a high proportion of musculoskeletal findings (81%). Neurodevelopmental phenotypes represent a significant burden and include intellectual disability (34%), autism spectrum disorder (38%), executive function deficits (46%), and graphomotor weakness (78%). Psychiatric illness manifests across the lifespan with schizophrenia prodrome (15%), psychosis (20%), anxiety disorders (40%) and ADHD (63%). On neurological exam study subjects displayed only mild or moderate motor difficulties.

**Conclusions:** By direct evaluation of 3q29 deletion study subjects, we document common features of the syndrome, including a high burden of neurodevelopmental and neuropsychiatric phenotypes. Evidence-based recommendations for evaluation, referral, and management are provided to help guide clinicians in the care of 3q29 deletion patients.

## Introduction

Individuals with 3q29 deletion syndrome (OMIM #609425) are hemizygous for a 1.6 Mb interval containing 21 protein coding genes^1^. The syndrome has a prevalence of ∼1 in 30,000 and is associated with reduced birth weight, failure to thrive, heart defects, intellectual disability, anxiety disorder, autism spectrum disorder, and schizophrenia^1–6^. Case reports have provided a rich source of data on individual patients and have illuminated many facets of the syndrome^2, 7–27^. However, phenotyping in case reports is rarely systematically applied, leading to bias in the curation of associated signs and symptoms^2^. Large cohorts of deidentified individuals referred for genetic testing^28–31^ or ascertained for a specific phenotype (such as autism spectrum disorder^32^ or schizophrenia^5, 33–35^) confirm the pathogenicity of the 3q29 deletion and provide evidence of increased risk for a single phenotype, but do not inform about the broader phenotypic spectrum. Systematic self-report of phenotypes from registries are also emerging, but are hampered by limitations of self-report data^3, 4^. For these reasons a comprehensive, unbiased characterization of the syndrome is needed, using gold standard instruments and direct evaluation of study subjects by trained experts^36^.

We sought to address these knowledge gaps in the clinical phenotype of 3q29 deletion syndrome. The Emory 3q29 Project unites an interdisciplinary team toward understanding the phenotypic spectrum, natural history, and molecular mechanisms of 3q29 deletion syndrome (http://genome.emory.edu/3q29/). As part of this project, our clinical team of investigators has directly and systematically evaluated 32 study subjects with the canonical 1.6 Mb 3q29 deletion using a standardized, transdiagnostic phenotyping protocol that includes gold-standard instruments^36^. These data have revealed new aspects of 3q29 deletion syndrome and provide guidance for the medical geneticist on recommended referral, follow-up, and management of patients.

## Materials and Methods

### Study subject eligibility

The study design, eligibility, recruitment criteria, and evaluation process have been previously described^36^. Briefly, individuals were recruited from the 3q29 registry housed at Emory University^3^ (3q29deletion.org). Inclusion criteria were: validated diagnosis of 3q29 deletion syndrome by documented chromosome microarray analysis where the deletion overlapped the canonical region (hg19, chr3:195725000-197350000) by >80%, and willingness and ability to travel to Atlanta GA. Exclusion criteria were: any 3q29 deletion with less than 80% overlap with the canonical region; non-fluency in English, and age younger than six years. One exception to the age criterion was made; a 4.85 year old who was part of a previously-described multiplex family was included in the study^27^. After an informed consent session with a staff scientist (MM), travel was arranged. This study was approved by the Emory Institutional Board (IRB000088012). Participant characteristics are summarized in Table 1.

**Table 1:** Study Subjects.

|  |  |
| --- | --- |
| Table 1: Study Subjects |  |
| Sex |  |
| Male | 20 (63%) |
| Female | 12 (37%) |
| Age - Mean (SD) | 14.5 (8.26) years |
| Age - Range | 4.85-39.12 years |
| Ethnicity (% non-Hispanic) | 31 (97%) |
| Race |  |
| White | 29 (91%) |
| More than 1 race | 3 (9%) |

### Evaluations

All study subjects were evaluated over two days at a designated clinical evaluation site. An in-person informed consent and assent procedure was repeated on the first study day.

Evaluations were conducted as follows (summarized in Table 2):

**Table 2:** Assessments and measures used in this study.

| Evaluation | Domain & Measures |
| --- | --- |
| Medical | Medical History<br>• Patient medical history by organ system<br>• Family medical history<br>• Family pedigree<br>Physical Examination<br>• Anthropomorphic Measures<br>• Physical examination by organ system<br>• Growth parameters (e.g., height, weight, head circumference) |
| Neurodevelopmental | Autism<br>• *Autism Diagnostic Interview -Revised* (ADI-R)<br>• *Autism Diagnostic Observation Schedule, 2<sup>nd</sup> ed* (ADOS-2)<br>Cognitive Ability & Adaptive Function<br>• Differential Abilities Scale, 2<sup>nd</sup> ed (DAS-II)<sup>a</sup><br>• Beery-Buktenica Visual Motor Integration Test-6<sup>th</sup> ed (VMI-6)<br>• *Behavioral Rating Inventory of Executive Functions, 2<sup>nd</sup> ed* (BRIEF-2)<sup>a</sup> and Adult version (BRIEF-A)<sup>b</sup><br>• Vineland Adaptive Behavior Scales, 3<sup>rd</sup> ed, Parent/Caregiver Interview Form<br>• Wechsler Abbreviated Scale of Intelligence, 2<sup>nd</sup> ed (WASI-II)<sup>b</sup> |
| Psychiatric | Anxiety<br>• *Anxiety Disorders Interview Schedule for DSM –IV (ADIS-IV) Child Version*<sup>a</sup><br>• *Structured Clinical Interview for DSM-V --Research Version* (SCID-5-RV) - Module F<sup>b, c</sup><br>Prodromal Symptoms & Psychosis<br>• Scheduled Interview for Prodromal Symptoms (SIPS)<br>• *Structured Clinical Interview for DSM-V --Research Version* (SCID-5-RV) - Module B/C<sup>b</sup><br>General Psychopathology<br>• *Kiddie Schedule for Affective Disorders and Schizophrenia* (K-SADS)<sup>a, c</sup><br>• *Structured Clinical Interview for DSM-V --Research Version* (SCID-5-RV) - Modules A, D, G, H, I, K<sup>b</sup> |
| Neurological | Gross and fine motor skill assessment |

<sup>a</sup> Administered to ages 6-18 years, <sup>b</sup> administered to ages 19+ years, <sup>c</sup> For some cases ages 19-22 years, the K-SADS was used to assess anxiety and psychopathology.

#### Medical History & Physical Examination

Parents completed a comprehensive medical history (Supplementary Information), which was reviewed during the visit with a trained medical geneticist (EB, RSR, MJG). The medical geneticist also completed a physical examination of the study subject. Photographs were taken of the front and profile of the face and any other physical characteristics of note. When possible, medical records were obtained to validate significant medical concerns.

Following the study visits, the medical geneticists met to discuss the results of the medical history and physical examination, identify relevant diagnoses by system, and classify each diagnosis using the Human Phenotype Ontology (HPO) terminology when available. HPO terminology provides a classification system for clinical abnormalities in humans (https://hpo.jax.org/app/). A detailed assessment of craniofacial characteristics was based upon physical examination notes and the photographs taken during the visit. Z-scores and percentiles for height, weight, and head circumference were determined according to *The Handbook of Physical Measurements*^*37*^ and were generated using the Face2Gene application.

#### Neurodevelopmental Evaluation

Table 2 summarizes a systematic battery used to evaluate neurodevelopmental phenotypes, which included assessments of cognitive ability (Differential Ability Scales, 2^nd^ ed.^38^ (DAS) for ages <18; Wechsler Abbreviated Scale of Intelligence^39^ (WASI) for ages > 18), adaptive behavior (Vineland Adaptive Behavior Scales 3rd ed.^40^), visual-motor integration (Beery-Buktenica Developmental Test of Visual-Motor Integration, 6^th^ ed.^41^ (VMI)), Autism spectrum disorder (Autism Diagnostic Interview, Revised^42^ (ADI-R); Autism Diagnostic Observation Schedule, 2^nd^ ed.^43^ (ADOS)), and executive function (Behavior Rating Inventory of Executive Functions^44^ (BRIEF-2 for ages <18, BRIEF-A for ages ≥ 18)).

Cognitive testing, VMI, and autism assessments were carried out by clinical psychologists (CS, CK, SW) early in the day to reduce fatigue. The Vineland and BRIEFs were completed by the parent or caretaker electronically via the publisher websites (Pearson Q-global and PARiconnect, respectively) either before or during the visit.

#### Psychiatric Evaluation

A systematic battery was also used to assess anxiety (Anxiety Disorders Interview Schedule for Children^45, 46^ (ADIS) in ages <18, Structured Clinical Interview for DSM-V Diagnosis, Research Version^47^ (SCID-5-RV) in ages 18+); general psychopathology (Schedule for Affective Disorders and Schizophrenia for School Aged Children^48^ (KSADS) in ages<18, SCID-5-RV^47^ in ages 18+); prodrome (Structured Interview for Prodromal symptoms^49, 50^ (SIPS)) and psychosis (SCID-5-RV). Prodrome and psychosis were evaluated in individuals age 8 years and older; at younger ages, developmentally appropriate magical thinking cannot be distinguished from true psychosis. Anxiety disorders in study subjects < 18 were evaluated with the ADIS, administered by a trained clinician (TLB). The KSADS, SCID-5-RV, and SIPS were administered by clinical psychology trainees (RE, DN, KG, KA, RG) supervised by a team clinical psychologist (EW). A qualitative assessment of global mental status and health was conducted by team psychiatrists (JFC, ME) to supplement impression and results from the formal assessments.

#### Neurological Exam

A subset of study subjects (n = 23) were evaluated by a pediatric neurologist (SK) for gross and fine motor phenotypes, including assessment of gait, heel to toe and stressed gait, fine finger movements, rapid alternating movements, as well as finger to nose and heel to shin tests. Performance on each task was rated using a 4-point scale of impairment severity: 0 (none/normal), 1 (mild), 2 (moderate), and 3 (severe). General clinical impressions were also noted. Exam results were recorded using a semi-structured data collection template; after the study visit data were entered into redcap.

## Results

### Participants

32 participants were consented and evaluated, including 4 participants from a single family^27^. Study subjects ranged in age from 4.8-39.1 years (mean age 14.5 years, median age 11.7 years); 62.5% (n = 20) were male. Study subjects were recruited from across the US, Canada, and the UK (Table 1).

### Medical history results

A summary of symptoms and diagnoses that occurred in at least 10% of study subjects evaluated is presented in Table 3. Supplemental table 1 incudes any symptoms or diagnosis reported in at least one study subject. The medical history review did not reveal any significant pre- or peri-natal findings that were generalizable across subjects.

**Table 3:** Frequency of symptoms and diagnoses reported in 10% or more of participants (N = 32)

| Symptoms/Diagnoses with Associated Procedure or | n | % |
| --- | --- | --- |

| Intervention |  |  |
| --- | --- | --- |
| **General** | **7** | **22%** |
| Fatigue | 7 | 22% |
| **HEENT** | **25** | **78%** |
| **Recurrent Infection addressed with surgery (any)** | **6** | **-** |
| *Cases requiring tonsillectomy* | 4 | - |
| *Cases requiring adenoidectomy* | 5 | - |
| **Eye (any)** | **19** | **59%** |
| Astigmatism | 5 | 16% |
| Myopia | 5 | 16% |
| Strabismus | 9 | 28% |
| *Cases with strabismus requiring surgery* | 3 | - |
| **Ear (any)** | **7** | **22%** |
| Recurrent ear infection | 7 | 22% |
| *Cases requiring myringotomy and tube placement* | 3 | - |
| **Nose (any)** | **8** | **25%** |
| Epistaxis | 7 | 22% |
| *Cases with epistaxis requiring surgery* | 2 | - |
| **Teeth (any)** | **13** | **41%** |
| Abnormal Number or Size of Teeth | 5 | 16% |
| Abnormal Dentition | 9 | 28% |
| **Cardiovascular** | **16** | **50%** |
| Structural | **15** | **47%** |
| Murmur | 7 | 22% |
| Complex congenital cardiovascular disease | 8 | 25% |
| *Cases requiring surgery* | 4 | - |
| **Respiratory** | **8** | **25%** |
| Asthma | 6 | 19% |
| **Sleep** | **10** | **31%** |
| Sleep Disturbance | 9 | 28% |
| **Gastrointestinal** | **26** | **81%** |
| Feeding Problems Beyond Infancy | 5 | 16% |
| Failure to Thrive Beyond Infancy | 13 | 41% |
| Constipation | 13 | 41% |
| Reflux | 16 | 50% |
| Feeding Problems in Infancy | 19 | 59% |
| **Renal/Genitourinary** | **9** | **28%** |
| Enuresis | 7 | 22% |
| **Integumentary/Dermatologic** | **11** | **34%** |
| Eczema | 4 | 13% |
| Keratosis Pilaris | 4 | 13% |
| **Allergy/immunology** | **9** | **28%** |
| Food Allergy | 4 | 13% |
| Seasonal Allergies | 5 | 16% |
| **Neurological (any)** | **18** | **56%** |
| Seizures (e.g., atonic, febrile, nocturnal) | 4 | 13% |
| Headache or Migraine | 5 | 16% |

## Review of systems

### Ear, nose, and throat (ENT)

78% of study subjects (n = 25) reported an ENT-related symptom or diagnosis in their medical history. 22% of subjects (n = 7) reported recurrent ear infections, and 3 required surgery. 59% (n = 19) had a symptom or diagnosis related to the eye. The most common ocular phenotype was strabismus (28%, n = 9), with 3 subjects requiring surgery. Vision problems encountered later in childhood included astigmatism (16%, n = 5) and myopia (16%, n = 5). Epistaxis was the most common nasal manifestation occurring in 22% of study subjects (n = 7), two of whom required surgical correction. Dental anomalies were reported in 41% (n = 13) and commonly included enamel hypoplasia with proclivity to caries and abnormalities in number and size of teeth.

### Cardiovascular

Structural cardiovascular disease is reported in 47% (n = 15) of patients. These included complex congenital cardiovascular disease (25%, n = 8) such as hypoplastic right heart (n = 1), patent ductus arteriosus (n = 2), pulmonary atresia (n = 2), pulmonary stenosis (n = 1), tricuspid stenosis (n = 1), ventricular septal defect (n = 2), and arterial-venous malformation (AVM) originating from the descending thoracic aorta (n = 1). Four individuals in this study (12.5%) required surgery to address their cardiovascular condition.

### Gastrointestinal

Gastrointestinal manifestations were noted in the majority (81%, n = 26) of patients. Among these, half of the patients (50%, n = 16) reported gastroesophageal reflux and 41% (n = 13) reported chronic constipation. Additionally, feeding difficulties in infancy were reported in more than half of patients (59%, n = 19) and persisted beyond infancy in 16% (n = 5). Only 9% (n = 3) of patients had failure to thrive in infancy, 2 of whom continued to have failure to thrive beyond infancy. In contrast, failure to thrive beyond infancy was reported in 41% (n = 13).

### Renal/Genitourinary

Enuresis was encountered in 25% (n = 8) of our cohort. Of these, seven had ongoing enuresis. The remaining study subject had a history of enuresis reported by parents during the medical history interview that was resolved at the time of this study.

### Respiratory, allergy, and immunological

Asthma was the most common respiratory manifestation, occurring in 19% (n = 6) of patients. Seasonal allergies were reported in 16% (n = 5) of subjects, two of whom also had asthma. Food allergies were noted in 13% (n = 4) of subject.

### Integumentary

13% (n = 4) of subjects reported a diagnosis of eczema. Keratosis pilaris was also reported in 13% (n = 4) of subjects.

### Sleep

Sleep disturbances were reported in at least 31% (n = 10) with 28% (n = 9) reporting difficulty initiating or maintaining sleep, and 1 person reporting sleep walking.

### Neurological

Seizures were reported in 13% (n = 4) of subjects. 16% (n = 5) of subjects reported experiencing headache or migraine.

### Medical History and Physical examination

Table 4 presents a summary of review of systems and physical findings (symptoms are not found on PE but reported by the patient) and diagnoses in at least 10% of study subjects evaluated. Supplemental table 2 incudes any symptoms or diagnosis found in at least one study subject.

**Table 4.**
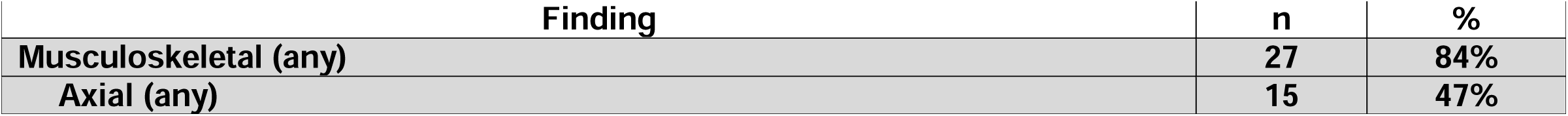
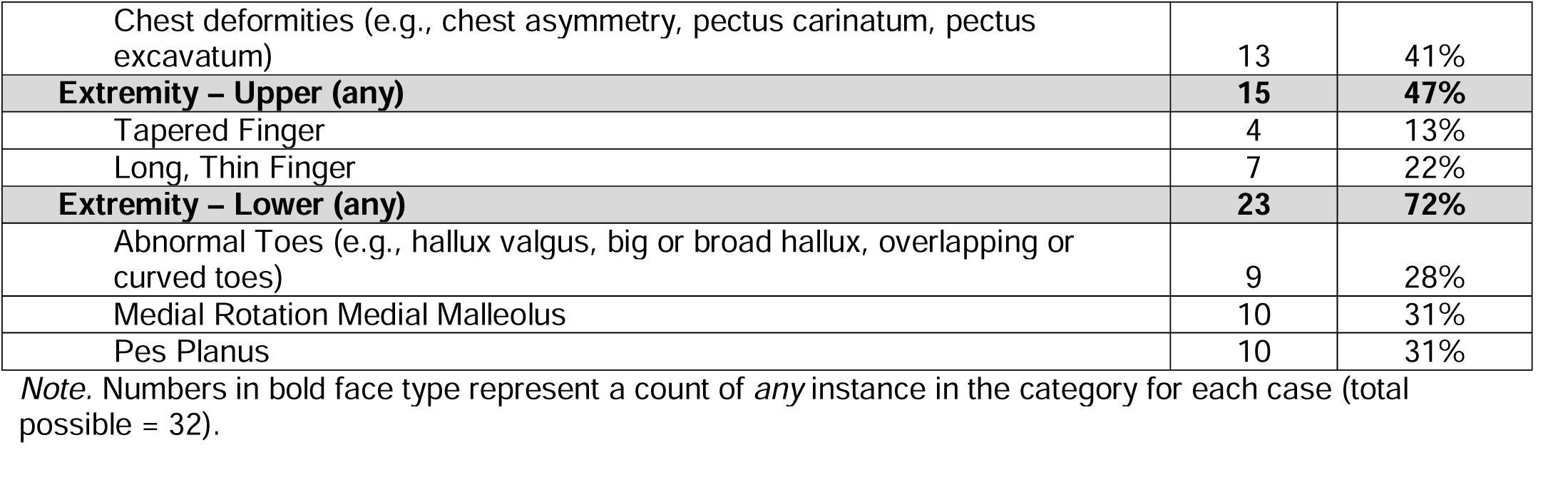
Frequency of Physical Exam Findings documented in 10% or more of participants.

### Growth parameters

Compared to age and sex norms, 25% of subjects (n = 8) had a weight −2 SD below the mean. These subjects ranged from 4-24 years and were evenly split between males and females. 16% of subjects (n = 5) were −2 SD below the mean for height. These subjects were 4 males and 1 female ranging in age from 4-39 years. 16% (n = 5) were −2 SD below the mean for head circumference. These subjects were 4 males and 1 female ranging from 4-12 years old. No subject had low weight, height, and head circumference. Most subjects had low measurements only in one parameter.

### Dysmorphic features

Subtle craniofacial dysmorphisms were noted among patients in the nasal region. These findings were identified by medical geneticists during the physical exam and quantified through next generation phenotyping technology through Face2Gene (FDNA, Inc., Boston, MA). See Mak et al (manuscript in preparation) for additional information.

### Musculoskeletal manifestations

On physical exam, 84% (n = 27) of study subjects had one or more musculoskeletal phenotypes. Chest deformities were noted in 41% (n = 13), and included pectus excavatum (25%, n = 8), pectus carinatum (9%, n = 3), and chest asymmetry (6%, n = 2). Manifestations in the upper extremities were seen in 47% of study subjects (n = 15). The most common findings were long, thin fingers (22%, n = 7) and tapered fingers (13%, n = 4). Lower extremity manifestations were seen in 72% of study subjects (n = 23). These included pes planus (31%, n = 10) or medial rotation of medial malleolus (31%, n = 10), with 22% having *both* pes planus and medial rotation of the medial malleolus. Abnormalities of the toes were also observed in 28% (n = 9) of study subjects, including abnormal hallux (13%, n = 4) commonly seen as broad hallux and hallux valgus, and abnormal non-hallux toes (19%, n = 6) frequently seen as curved, short, or overlapping toes.

Within the other systems examined, individual symptoms or diagnoses were noted (see Supplemental Table 2), but no single finding was frequent enough to be generalizable across study subjects.

## Neurodevelopmental Phenotypes

Rates of neurodevelopmental and psychiatric diagnoses are shown in Figure 1; quantitative mean and median scores with ranges are shown in Table 5.

**Table 5.** Mean and median scores on neurodevelopmental instruments.

| Domain | Mean | Median | Range |
| --- | --- | --- | --- |
| Cognitive, standard score (GCA or FSIQ) | 73.0 | 75.5 | 99-40 |
| Adaptive behavior, standard score | 73.9 | 70.5 | 48-110 |
| Executive Function, T-score | 68.3 | 69.0 | 45-88 |
| Visual Motor Integration, standard score | 69.5 | 67 | 45-103 |

*Standard Scores* have an expected population mean of 100 and a standard deviation of 15; scores *below* 70 (2 SD from the expected mean) are considered to denote a clinically significant weakness. *T-scores* have an expected population mean of 50 and a standard deviation of 10; scores *above* 70 (2 SD from the expected mean) are considered to denote a clinically significant weakness.

**Figure 1:**
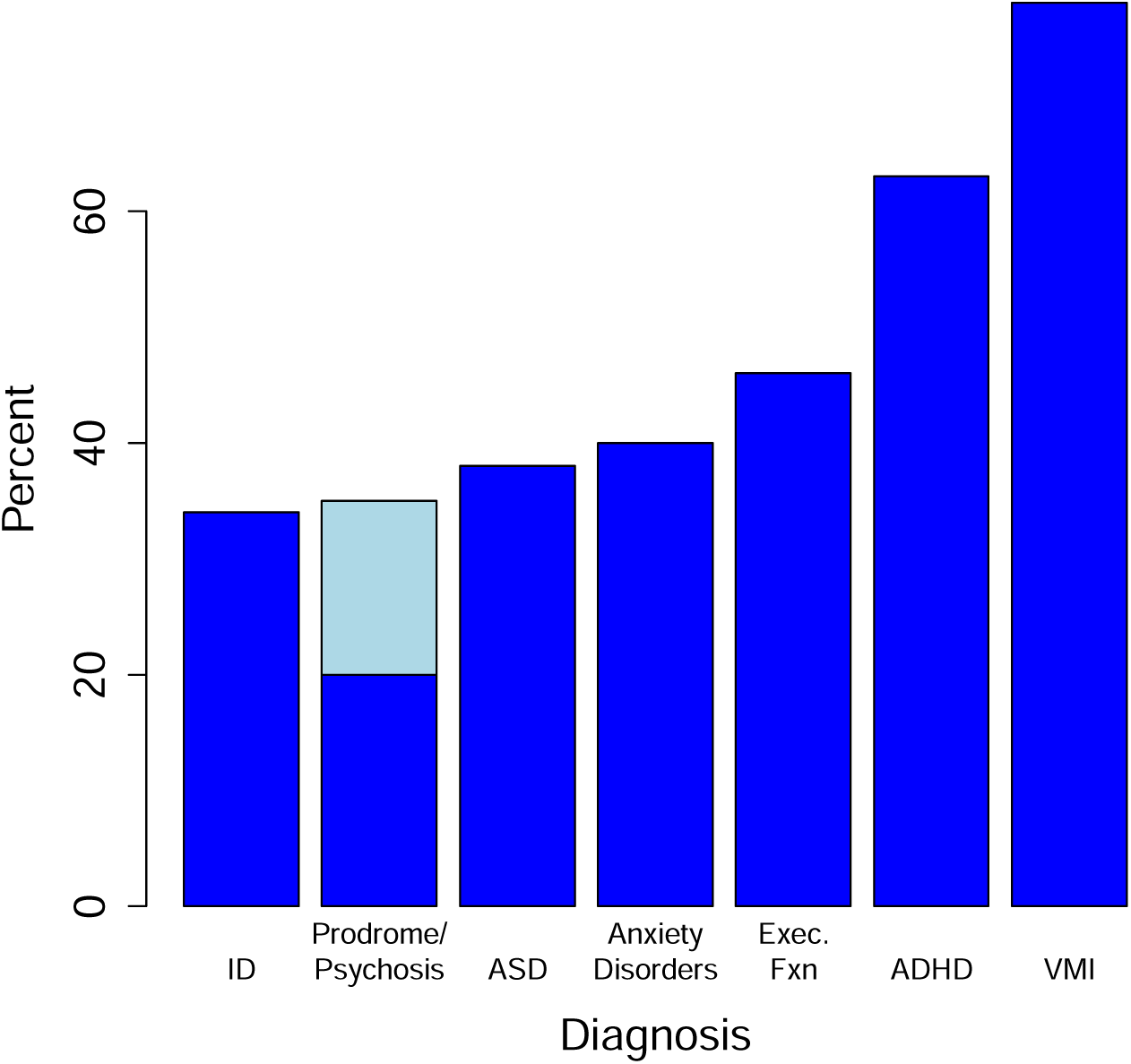
Percent of 3q29 deletion participants who qualify for neurodevelopmental and neuropsychiatric diagnoses after direct evaluation by our team. 19% of study subjects qualified for a diagnosis of psychosis; 15% had features of schizophrenia prodrome (light blue). ID, intellectual disability; ASD, autism spectrum disorder; ADHD, attention-deficit hyperactivity disorder; VMI, visual-motor integration.

### Cognitive Ability and Adaptive Function

Mean IQ score (FSIQ) was 73.0 (median 75.5, range 40-99). Mean adaptive function as measured with the Vineland was 73.9 (median 70.1, range 48-107). Individuals were considered to have intellectual disability if both GCA and adaptive behavior scores were >2 SD below the expected mean (> 70 on both evaluations), consistent with DSM-V criteria. By this metric, 11 individuals (34%) qualified for a diagnosis of ID.

### Graphomotor Weakness

78% of study subjects were found to have clinically significant graphomotor weakness, defined as a score < 2 SD below the expected mean. Examination of subtest scores indicated greater weakness in motor coordination than visual perception.

### Executive Function

47% of study subjects (n = 15) were found to have clinically significant deficits in Executive Function, with T-scores that were ≥ 2 SD above the mean.

### Autism

Autism was assessed using the ADOS and ADI-R; these instruments were used to inform the administering clinician’s best estimate diagnosis. Of 32 individuals evaluated, 12 (37.5%) met DSM-V criteria for a diagnosis of Autism Spectrum Disorder. Nine of 20 males evaluated (45%) qualified for a diagnosis of autism, a 19.5-fold excess as compared to the general population rate of 1 in 42 males (2.3%)^51^. Three out of 12 females evaluated (25%) also qualified for a diagnosis of autism, a 50-fold excess compared to the general population rate of ASD in females (1 in 189, 0.5%)^51^. These data confirm there is a high risk for autism associated with the 3q29 deletion in both males and females.

## Psychiatric Evaluation

### Anxiety Disorders

40% (n = 13) of study subjects qualified for at least one anxiety disorder; 18% (n=6) had more than one anxiety disorder diagnosis. Anxiety disorders included generalized anxiety disorder (22%, n = 7), specific phobia (19%, n = 6), separation anxiety (12.5%, n = 4) and social anxiety disorder (6%, n = 2).

### Prodromal Symptoms & Psychosis

Of 21 individuals who were evaluated, 19% (n = 4) qualified for diagnosis of a psychotic disorder. Three additional individuals (14%) were found to exhibit features consistent with the schizophrenia prodrome. These data are consistent with prior reports establishing the 3q29 deletion as a risk factor for schizophrenia^5, 6, 33–35^.

### General Psychopathology

63% (n = 20) of individuals evaluated qualified for a diagnosis of attention-deficit hyperactivity disorder (ADHD). 19 of these individuals could be further classified into subtypes: ADHD-inattentive type (n = 10), ADHD-combined type (n = 8), and ADHD-Hyperactive/impulsive (n = 1). One additional individual did not fit criteria for any subtype and was diagnosed as “other specified ADHD.” The high rate of ADHD in 3q29 deletion study subjects is consistent with deficits in executive function, as reported above. No other psychiatric diagnosis was present in great than 10% of study subjects. Two people met criteria for intermittent explosive disorder; two people met criteria for obsessive compulsive disorder; two people met criteria for major depressive disorder. For each of the following diagnoses, one person met criteria: oppositional defiant disorder, conduct disorder, Bipolar I disorder.

## Neurological Exam

A subset of study subjects (n = 23) were evaluated by a trained pediatric neurologist for motor phenotypes associated with cerebellar dysfunction. Motor phenotypes were identified, but generally were mild (range 30-47%) or moderate (range 4-48%, Table 6). Only one person displayed a severe cerebellar motor phenotype, in rapid alternating movement.

**Table 6:** Results of Neurological Exam for 23 individuals with 3q29 deletion syndrome.

|  | Fine Finger Movement | Rapid Alternating Movement | Heel to Shin Task | Finger to Nose Task | Tandem Walk |
| --- | --- | --- | --- | --- | --- |
| normal/score 0 (%) | 6 (26%) | 4 (17%) | 6 (32%) | 14 (61%) | 11 (55%) |
| Mild/score 1 (%) | 9 (39%) | 7 (30%) | 9 (47%) | 8 (35%) | 6 (30%) |
| moderate/score 2 (%) | 8 (35%) | 11 (48%) | 4 (21%) | 1 (4%) | 3 (15%) |
| severe/score 3 (%) | 0 (0%) | 1 (4%) | 0 (0%) | 0 (0%) | 0 (0%) |
| not normal (%) | 17 (74%) | 19 (83%) | 13 (68%) | 9 (39%) | 9 (45%) |

## Discussion

Here we report the first comprehensive description of 3q29 deletion syndrome, by direct systematic evaluation of 32 study subjects using a defined protocol and gold-standard instruments. While nearly all major systems are affected in 3q29 deletion syndrome, the ocular, dental, cardiovascular, gastrointestinal, renal, musculoskeletal and neurologic systems are affected with the greatest frequency and should be prioritized for evaluation and follow-up. There is also a high burden of neurodevelopmental and neuropsychiatric illness, requiring intervention and support across the lifespan. Our findings motivate recommendations for clinical care, described in detail below and summarized in Table 7.

**Table 7:**
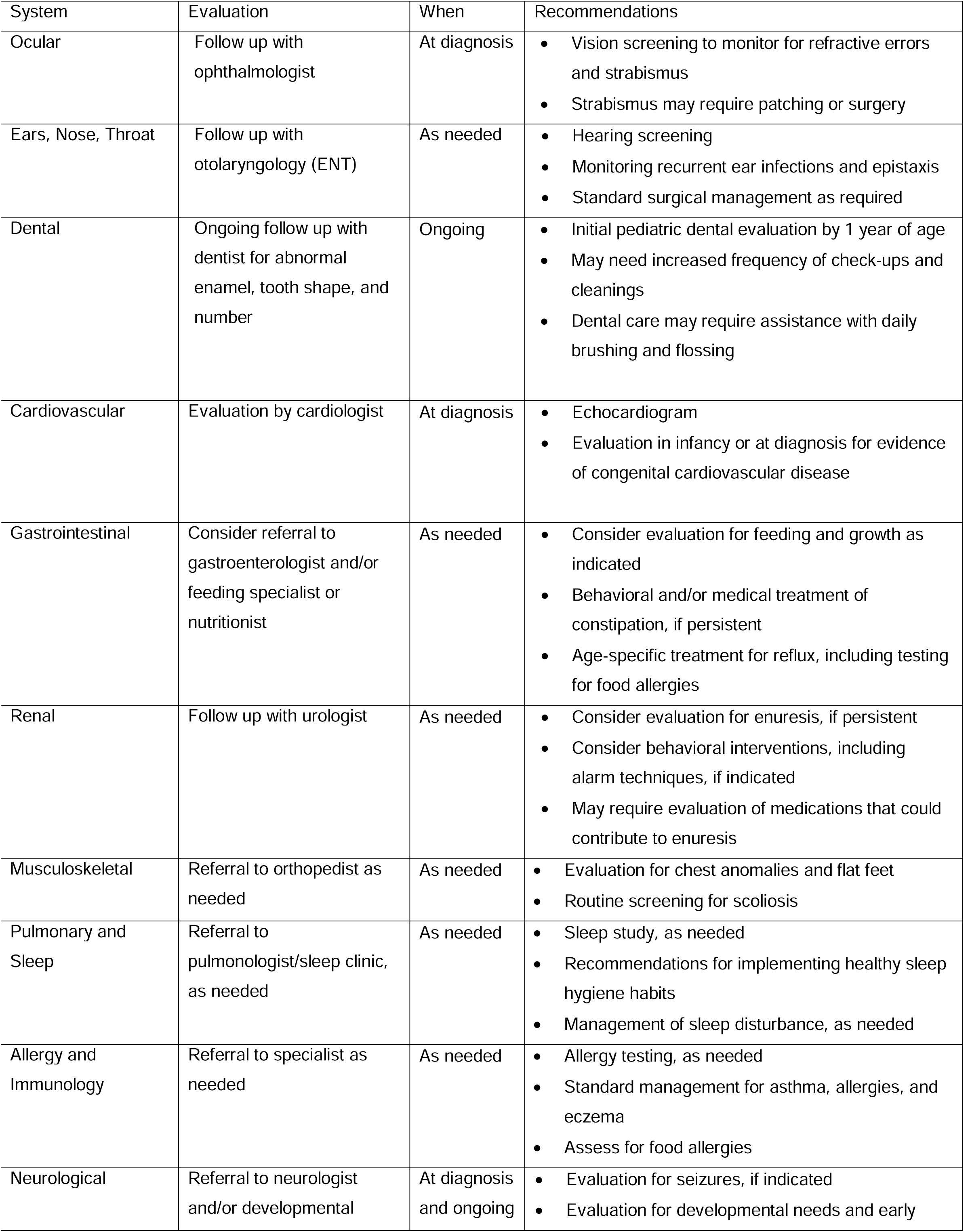
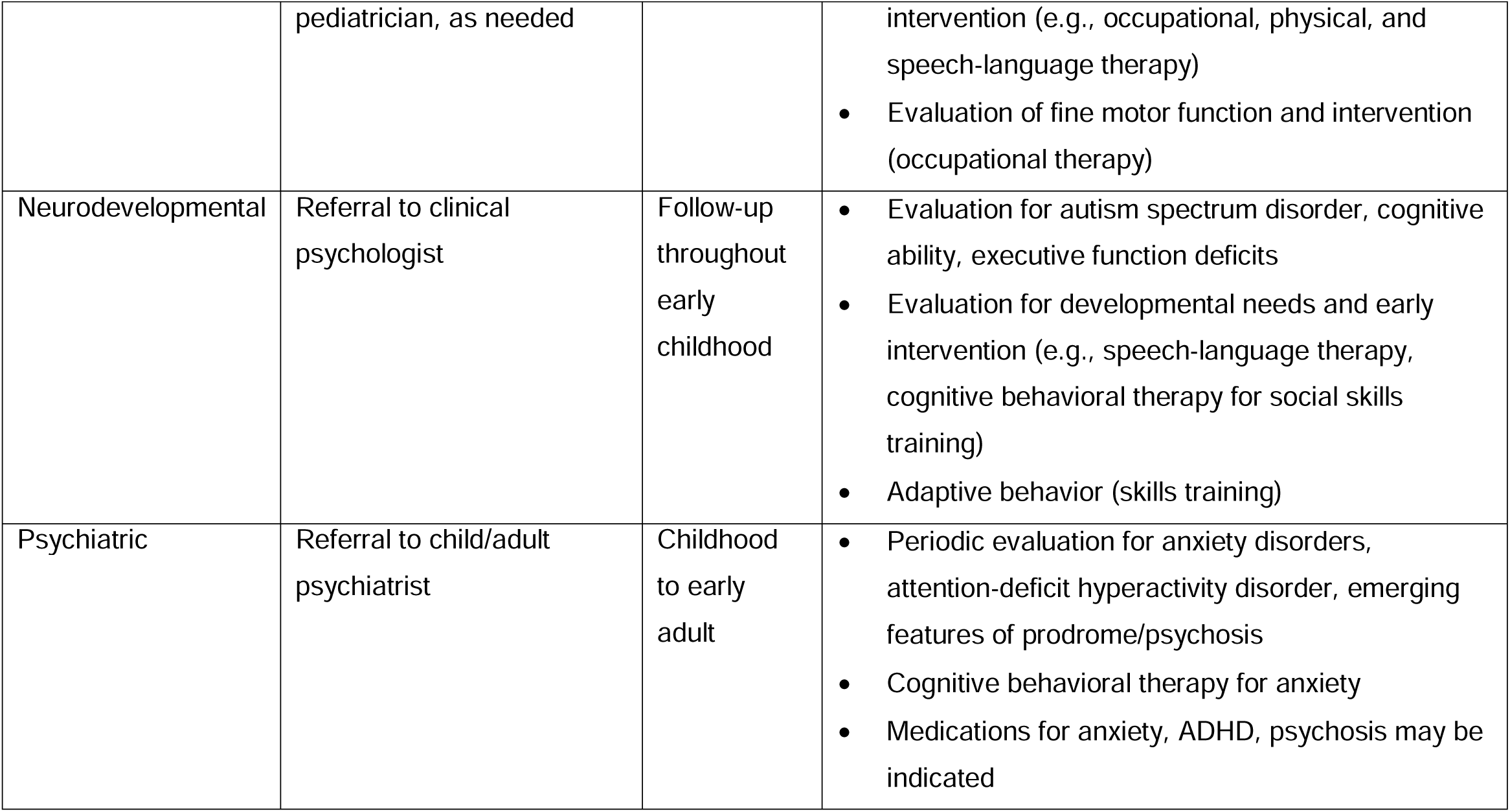
Recommendations for clinical care for individuals with 3q29 deletion syndrome.

| System | Evaluation | When | Recommendations |
| --- | --- | --- | --- |
| Ocular | Follow up with ophthalmologist | At diagnosis | • Vision screening to monitor for refractive errors and strabismus<br>• Strabismus may require patching or surgery |
| Ears, Nose, Throat | Follow up with otolaryngology (ENT) | As needed | • Hearing screening<br>• Monitoring recurrent ear infections and epistaxis<br>• Standard surgical management as required |
| Dental | Ongoing follow up with dentist for abnormal enamel, tooth shape, and number | Ongoing | • Initial pediatric dental evaluation by 1 year of age<br>• May need increased frequency of check-ups and cleanings<br>• Dental care may require assistance with daily brushing and flossing |
| Cardiovascular | Evaluation by cardiologist | At diagnosis | • Echocardiogram<br>• Evaluation in infancy or at diagnosis for evidence of congenital cardiovascular disease |
| Gastrointestinal | Consider referral to gastroenterologist and/or feeding specialist or nutritionist | As needed | • Consider evaluation for feeding and growth as indicated<br>• Behavioral and/or medical treatment of constipation, if persistent<br>• Age-specific treatment for reflux, including testing for food allergies |
| Renal | Follow up with urologist | As needed | • Consider evaluation for enuresis, if persistent<br>• Consider behavioral interventions, including alarm techniques, if indicated<br>• May require evaluation of medications that could contribute to enuresis |
| Musculoskeletal | Referral to orthopedist as needed | As needed | • Evaluation for chest anomalies and flat feet<br>• Routine screening for scoliosis |
| Pulmonary and Sleep | Referral to pulmonologist/sleep clinic, as needed | As needed | • Sleep study, as needed<br>• Recommendations for implementing healthy sleep hygiene habits<br>• Management of sleep disturbance, as needed |
| Allergy and Immunology | Referral to specialist as needed | As needed | • Allergy testing, as needed<br>• Standard management for asthma, allergies, and eczema<br>• Assess for food allergies |
| Neurological | Referral to neurologist and/or developmental | At diagnosis and ongoing | • Evaluation for seizures, if indicated<br>• Evaluation for developmental needs and early |
|  | pediatrician, as needed |  | intervention (e.g., occupational, physical, and speech-language therapy)<br>• Evaluation of fine motor function and intervention (occupational therapy) |
| Neurodevelopmental | Referral to clinical psychologist | Follow-up throughout early childhood | • Evaluation for autism spectrum disorder, cognitive ability, executive function deficits<br>• Evaluation for developmental needs and early intervention (e.g., speech-language therapy, cognitive behavioral therapy for social skills training)<br>• Adaptive behavior (skills training) |
| Psychiatric | Referral to child/adult psychiatrist | Childhood to early adult | • Periodic evaluation for anxiety disorders, attention-deficit hyperactivity disorder, emerging features of prodrome/psychosis<br>• Cognitive behavioral therapy for anxiety<br>• Medications for anxiety, ADHD, psychosis may be indicated |

Almost 60% of subjects reported ocular manifestations, the most frequently occurring ocular manifestations were strabismus and refractive errors, including myopia and astigmatism. In some cases, the strabismus required surgical repair. The high rate of ocular manifestations is consistent with previous reports^2^ and suggests the importance of involving a pediatric ophthalmologist at time of diagnosis for standard treatment of refractive errors and strabismus.

Similar to prior reports^2, 3, 7^, recurrent ear infections were common among study subjects and sometimes required myringotomy tubes. Epistaxis has not been previously reported but was commonly seen in a little over 20% of subjects without a concurrent history of nose picking. Two of the seven cases required surgery, one of which had developed anemia. The high rates of ear infections and epistaxis suggest a low threshold for involving ENT in care. When taken together with increased risk for speech-language disorders, these results further highlight the importance of hearing screening to monitor for conductive hearing loss and associated disruptions to speech-language development.

Consistent with reports of dental anomalies in the literature^2, 3, 7^, parents of 41% of subjects in our study reported a broad spectrum of dental anomalies, such as abnormal tooth shape, size, and number, diastema, enamel hypoplasia, and frequent cavities. In some cases, extensive surgical intervention was required, including up to 5 root canals in a single individual. Cavities occurred despite parent reported good oral hygiene. Notably, our study and others^2, 3, 16^ report dental anomalies along with gastroesophageal reflux, which may contribute to dental problems. Regardless, the high frequency of dental problems among individuals with 3q29 deletion highlights the need for early and ongoing dental care by a pediatric dentist, which may include increased frequency of routine check-ups and cleanings along with education in dental hygiene.

Previous literature draws attention to elevated rates of heart defects in individuals with 3q29 deletion^2, 3, 16^. Our findings confirm this increased risk. Structural cardiovascular manifestations were reported in almost half of subjects. Almost all manifestations were heart defects ranging in severity from PDA to hypoplastic right heart syndrome. No single common congenital heart defect was observed. One subject had a large AVM originating from the descending aorta, which was incidentally found on imaging for back pain. Of note, however, is that this subject also had an additional loss of 188 kb in 7q31.31 of uncertain significance that included only 1 OMIM gene, KCND2. To date AVMs have not typically been associated with *KCND2* or described in 3q29, thus it is unclear whether this AVM is related to 3q29. Additional genetic testing was not performed. Regardless, it is important to note that half of subjects with a cardiovascular manifestation required surgical repair, suggesting the need for echocardiogram at diagnosis, if not previously done.

Gastrointestinal manifestations occurred in 81% of subjects and are noted as early as infancy. Feeding difficulties in infancy, manifesting as problems with latching to the breast and sucking, were sometimes associated with failure to gain weight and were common presenting signs. Feeding difficulties, coupled with reflux and constipation, and less frequently, dysphagia and esophageal dysmotility, persisted beyond infancy. Three subjects needed gastrostomy tubes beyond infancy due to restrictive food preferences and failure to thrive. It is unclear how much feeding problems may contribute to the observed smaller height and lower weight as documented in growth parameters (discussed subsequently) and/or whether these subjects are constitutionally smaller.

Occurring in 22% of subjects, enuresis was the most common renal/genitourinary manifestation reported. The subjects with a diagnosis of enuresis ranged in age from 6 to 16.9 years, suggesting that enuresis continues beyond the age in which it often resolves spontaneously. Enuresis is typically diagnosed more commonly among males than females in the general population.

However, this sex difference was not apparent among our subjects, 4 of whom were male and 3 of whom were female, which supports the notion that the 3q29 deletion contributes to enuresis. Indeed, prior case studies report a 15-year-old male^2^ and an 8-year-old female^13^ with urinary voiding dysfunction. Although varied by age and gender, the frequency of enuresis is reported to be 2.8% for children ages 6-14 years in the general population, indicating an approximately 9-fold enrichment among our study subjects and may require behavioral or pharmacological intervention.

A constellation of respiratory, allergy, and immunological symptoms were reported among study subjects. We observed 19% of our study subjects had asthma, 16% had seasonal allergies, and 13% had food allergies. The rate of asthma and food allergies are higher than the rates reported in the general population^52, 53^. Although the rates of seasonal allergy did not exceed the rate of 10-20% in the general population^54^, these results taken together suggest a tendency to atopy. Moreover, common skin manifestations reported among study subjects included eczema and keratosis pilaris, both of which are also associated with atopy. Early evaluation and routine follow-up with a pediatric pulmonologist or allergist may be important for ongoing care.

Although not previously reported in the literature, sleep disturbances were reported in 31% of subjects and ranged from difficulty initiating or maintaining sleep to reporting sleep walking and sleep apnea. Sleep disturbances can co-occur with other diagnoses, such as enuresis^55^. Interestingly, half of the subjects in our cohort with enuresis also had a sleep disturbance. Diagnosis and management of sleep disorders, particularly among subjects with other co-morbidities, may lead to improvement in multiple areas.

Based upon growth parameters, preliminary analyses were conducted to explore the effect of sex and age on height and weight. Results for height and weight suggest there may be a sex by age interaction. Male children are only slightly shorter than the mean, but by adulthood are shorter as compared to the mean than they were as children. In contrast, females move in the opposite direction: they are short as children but move to the population average as adults. In adulthood, males with 3q29 deletion are shorter (relative to expectations) than females (p value 0.01). At young ages, females weight less than expected but catch up in adulthood. Males move in the opposite direction: They are closer to the mean as children and by adulthood weigh less than expected. Only an age effect was noted for head circumference. Children with 3q29 deletion (males and females) have an FOC that is well below the population mean, but by adulthood this effect is ameliorated.

There is a substantial burden of disability contributed by neurodevelopmental and psychiatric manifestations. Cognitive disability is present but mild to moderate in most of the study subjects we evaluated. However, there is likely a cumulative impact of diminished cognitive ability alongside multiple comorbidities, including autism spectrum disorder, executive function deficits, anxiety disorders, and ADHD, that impair functioning and are a threat to successful social and occupational functioning. The high rate of graphomotor weakness indicates that writing and other fine motor tasks may be effortful and introduce challenges in academic settings. Occupational therapy may be helpful. Creative classroom solutions to alleviate the burden of writing tasks may decrease stress in these settings.

The high rate of ADHD, anxiety disorders, and schizophrenia/psychosis indicate that a child/adolescent psychiatrist should be part of the medical team, with periodic evaluation throughout development to detect emerging symptoms. Early intervention with cognitive behavioral therapy may be effective for anxiety disorders. Anxiety disorders, ADHD, and psychosis may require control with pharmaceutical agents. Because of the risk of psychosis associated with the 3q29 deletion, it is recommended that stimulant use for ADHD treatment be avoided if possible.

Limitations of this study include the small sample size, though we note that 32 study subjects is a relatively large sample size given the low frequency (1 in 30,000) of 3q29 deletion syndrome.

However, this sample size allows us to describe only the most common manifestations of the syndrome; a larger sample size mat reveal lower frequency symptoms Because our study required travel to Atlanta GA, this may have introduced some ascertainment bias toward subjects who were healthy enough to travel; subjects with behavioral challenges may not have participated. Because this sample is relatively young (average age 14.5 years) we have limited ability to describe later onset symptoms of the syndrome. Future directions will include a larger sample size and longitudinal evaluation.

In conclusion, many of the medical, neurodevelopmental, and psychiatric findings, although frequent, are nonspecific. Many symptoms noted require medical intervention, but do not necessarily signal the need for genetic testing. As a result, in individuals with 3q29 deletion syndrome, the need for genetic testing may be easily overlooked in the absence of other major congenital anomalies, thus getting to diagnosis remains a challenge. Taking into account the whole constellation of symptoms, including developmental and neuropsychiatric manifestations, are necessary to ensure patient access to genetic testing. Once 3q29 deletion syndrome is diagnosed, the findings and recommendations in the current study provide clinicians and families a road map toward effective strategies for care and treatment for individuals with 3q29 deletion syndrome.

## Data Availability

Data from this manuscript are available in NDAR: https://nda.nih.gov/about.html

## Acknowledgements

3q29 project members

NIH grants R01 MH110701 and R01 MH118534.

The 3q29 community

The Marcus Autism Center

Please note that any publication that results from a project utilizing REDCap should cite grant support (UL1 TR000424))

